# No evidence of clinical efficacy of hydroxychloroquine in patients hospitalised for COVID-19 infection and requiring oxygen: results of a study using routinely collected data to emulate a target trial

**DOI:** 10.1101/2020.04.10.20060699

**Authors:** Matthieu Mahévas, Viet-Thi Tran, Mathilde Roumier, Amélie Chabrol, Romain Paule, Constance Guillaud, Sébastien Gallien, Raphael Lepeule, Tali-Anne Szwebel, Xavier Lescure, Frédéric Schlemmer, Marie Matignon, Medhi Khellaf, Etienne Crickx, Benjamin Terrier, Caroline Morbieu, Paul Legendre, Julien Dang, Yoland Schoindre, Jean-Michel Pawlotski, Marc Michel, Elodie Perrodeau, Nicolas Carlier, Nicolas Roche, Victoire de Lastours, Luc Mouthon, Etienne Audureau, Philippe Ravaud, Bertrand Godeau, Nathalie Costedoat-Chalumeau

## Abstract

**Background:** Treatments are urgently needed to prevent respiratory failure and deaths from coronavirus disease 2019 (COVID-19). Hydroxychloroquine (HCQ) has received worldwide attention because of positive results from small studies.

**Methods:** We used data collected from routine care of all adults in 4 French hospitals with documented SARS-CoV-2 pneumonia and requiring oxygen ≥ 2 L/min to emulate a target trial aimed at assessing the effectiveness of HCQ at 600 mg/day. The composite primary endpoint was transfer to intensive care unit (ICU) within 7 days from inclusion and/or death from any cause. Analyses were adjusted for confounding factors by inverse probability of treatment weighting.

**Results:** This study included 181 patients with SARS-CoV-2 pneumonia; 84 received HCQ within 48 hours of admission (HCQ group) and 97 did not (no-HCQ group). Initial severity was well balanced between the groups. In the weighted analysis, 20.2% patients in the HCQ group were transferred to the ICU or died within 7 days vs 22.1% in the no-HCQ group (16 vs 21 events, relative risk [RR] 0.91, 95% CI 0.47–1.80). In the HCQ group, 2.8% of the patients died within 7 days vs 4.6% in the no-HCQ group (3 vs 4 events, RR 0.61, 95% CI 0.13–2.89), and 27.4% and 24.1%, respectively, developed acute respiratory distress syndrome within 7 days (24 vs 23 events, RR 1.14, 95% CI 0.65–2.00). Eight patients receiving HCQ (9.5%) experienced electrocardiogram modifications requiring HCQ discontinuation.

**Interpretation:** These results do not support the use of HCQ in patients hospitalised for documented SARS-CoV-2-positive hypoxic pneumonia.

## Introduction

The WHO-declared pandemic of severe acute respiratory syndrome coronavirus 2 (SARS-CoV-2) is causing fatal pneumonia from coronavirus disease 2019 (COVID-19). Treatments are urgently needed to prevent hypoxemic respiratory failure and death.^1^ Because an in vitro study documented potential activity by hydroxychloroquine (HCQ) on SARS-CoV-2^2^ and small studies have released controversial results, HCQ has received intense worldwide attention. Its effectiveness for COVID-19 is highly contentious.^3^ One uncontrolled French study of 26 hospitalised patients with SARS-CoV-2 PCR on a nasopharyngeal swab suggested that HCQ, at a dose of 600 mg/day, decreased SARS-CoV-2 shedding and that the combination with azithromycin had further efficacy.^4^ However, another uncontrolled French study found no evidence of antiviral clearance with HCQ and azithromycin in 11 hospitalised patients.^5^ From a clinical perspective, a recent study randomised 62 patients in two parallel groups, one control group and one receiving HCQ treatment (400 mg/d for 5 days), and found a reduction of time to clinical recovery.^6^ These patients, however, were not severely ill, the clinical endpoints were not clearly defined, and there was no stratification for comorbidities associated with a poor outcome.^6^

Based on these results, the negligible cost and known safety profile of HCQ, this drug has been considered to be potentially useful in patients with SARS-CoV-2, has attracted enormous attention in social and mass media and has received FDA approval for severe cases.^7^ On the other hand, fears of a shortage of this essential treatment for patients with rheumatic diseases including systemic lupus erythematous (SLE) have recently increased, and questions about its safety in patients with COVID-19 have been raised.^3^

Due to the lack of unbiased data and the urgency of confirming, if possible, the clinical efficacy of HCQ in this situation, we designed an emulated trial using observational data collected in a real-world setting in patients hospitalised for COVID-19 infection requiring oxygen. The primary aim was to evaluate the clinical effectiveness of oral HCQ at a daily dose of 600 mg in preventing admission to the intensive care unit (ICU) and/or death by any cause; secondary aims were to assess its effectiveness in preventing acute respiratory distress syndrome (ARDS).

## Methods

### Study Design

We used data collected from routine care to emulate a target trial aimed at assessing the effectiveness of HCQ for patients hospitalised with a COVID-19 infection and requiring oxygen.^8^

### Study population

To assess patient eligibility, physicians screened the electronic health records of all patients hospitalised between March 17 and 31, 2020, in four French tertiary care centres providing care to patients with COVID-19 pneumonia. Adult patients were eligible in this study if they were aged between 18 years and 80 years, had PCR-confirmed SARS-CoV-2 infection, and required oxygen by mask or nasal prongs (corresponding to a WHO progression score of 5).

Exclusion criteria were 1) the presence of a contraindication to HCQ at 600 mg daily (including patients under dialysis); 2) the start of HCQ before admission to the hospital; 3) treatment with another experimental drug for COVID-19 (tocilizumab, lopinavir-ritonavir, or remdesivir) within 48 hours after admission; 4) organ failure requiring immediate admission to the ICU or continuous care unit (CCU); 5) ARDS at admission (defined by the need for non-invasive ventilation with provision of positive airway pressure or invasive mechanical ventilation);^9^ 6) discharge from the ICU to standard care; 7) decision to limit and stop active therapeutics made at admission); and 8) opposition to data collection by the patient or her/his legal representative.

The study was performed in accordance with the declaration of Helsinki, as amended, and received approval by the appropriate IRB (number 2020_060, Paris XII University, AP-HP).

### Treatment strategies

We compared two treatment strategies: initiation of HCQ at a daily dose of 600 mg in the first 48 hours after hospitalisation (HCQ group) and the absence of HCQ initiation during this two-day period (no-HCQ or control group).

### Start and end of follow-up

To avoid time-dependent bias, the start of follow-up (baseline or time zero) for each individual was the time of his or her hospital admission. The end of follow-up was death, discharge home, or day 7 (D7) after hospitalisation.

### Outcomes

The primary outcome was a composite: transfer to the ICU within 7 days of inclusion and/or death from any cause. Secondary outcomes were all-cause mortality at day 7 and the occurrence of ARDS. For patients transferred to another hospital, physicians were contacted to obtain outcome data; if this was unsuccessful, these outcome data were considered missing. Before HCQ initiation and 3 to 5 days afterwards, all patients receiving HCQ had QT prolongation assessed by a 12-lead electrocardiogram (ECG) and corrected for heart rate by Bazett’s or Fredericia’s formula.

### Statistical analysis

An inverse probability of treatment weighting (IPTW) approach was used to “emulate” randomisation and balance the differences in baseline variables between treatment groups.^10,11^ A non-parsimonious multivariable logistic regression model was constructed to estimate each patient’s probability of receiving HCQ given their baseline covariates (i.e., the propensity score). Variables of the propensity score (PS) model were planned and prespecified before outcome analyses and included 1) age; 2) gender; 3) comorbidities (presence of chronic respiratory insufficiency under oxygen therapy or asthma or cystic fibrosis or any chronic respiratory pathology likely to decompensate during a viral infection; heart failure (NYHA III or IV); chronic kidney disease; liver cirrhosis with a Child-Pugh stage B or more; personal history of cardiovascular disease [hypertension, stroke, coronary artery disease, cardiac surgery]; insulin-dependent diabetes mellitus, or diabetic microangiopathy or macroangiopathy; treatment by immunosuppressive drugs, including anticancer chemotherapy; uncontrolled HIV infection or HIV infection with CD4 cell counts < 200/µl; or a haematological malignancy); 4) BMI (≥30 kg/m^2^ or not); 5) third trimester of pregnancy; 6) treatment by angiotensin-converting enzyme inhibitors (ACEIs) or angiotensin receptor blockers (ARBs);^12^ 7) time since symptom onset; and 8) severity of condition at admission (percentage of lung affected [≥50% or not]; presence of confusion; respiratory frequency; oxygen saturation without oxygen; oxygen flow; systolic blood pressure; and C-reactive protein (CRP) (≥40 mg/L or not)). Non-linear effects of continuous variables were considered in the model by using fractional polynomials. Standardised differences were examined to assess balance, with a threshold of 10% designated to indicate clinically meaningful imbalance.^13^ Outcomes were analysed based on the IPTW estimate of the relative risk (RR), with its 95% confidence interval (CI) derived from the variance estimator accounting for the estimated propensity score.^14^ We also examined outcomes in the subgroup of patients with a better prognosis at admission, estimated by a qSOFA < 2.^14,15^

Several sensitivity analyses were conducted to assess the robustness of findings. First, we analysed the unweighted sample. Second, we performed a trimmed analysis that was truncated at 5% of the extreme weights. Finally, we compared our results with an analysis that excluded patients who received HCQ later during their follow-up (i.e., more than 48 h after admission).

We had no missing data for either transfer to ICU or death, and only two cases missing data about ARDS. We chose not impute missing data. All statistical analyses were performed with the R statistical package version 3.6.1 or later (The R Foundation for Statistical Computing, https://www.R-project.org/).

## Results

### Patients

Among the 181 patients eligible for analysis, 84 received HCQ within 48 hours of admission and 97 did not (although 8 of them did receive HCQ later on). The median age of patients was 60 years (interquartile range [IQR], 52 to 68 years), and 71.1% were men. All comorbidities were less frequent in the HCQ group. The median delay between symptom onset and admission to hospital was 7 days (IQR, 5 to 10 days). Overall, initial severity was well balanced between the groups, except for confusion at admission (0 in the HCQ group vs 6 [6.2%] in the no-HCQ group) (**Table 1**). Further, of patients in the HCQ group, 17 (20%) received concomitant azithromycin, and 64 (76%) received concomitant amoxicillin and clavulanic acid.

**Table 1:** Demographic and clinical characteristics of the patients at baseline. Abbreviations: IQR, interquartile range; HCQ, hydroxychloroquine; NYHA, New York Heart Association; BMI, body mass index; ACEIs, angiotensin-converting enzyme inhibitors; ARBs, angiotensin receptor blockers.

| Characteristic | Total<br>(n=181) | HCQ<br>(n=84) | No-HCQ<br>(n=97) |
| --- | --- | --- | --- |
| **Demographic and clinical data** |  |  |  |
| Age, median (IQR) -year | 60 [52 – 68] | 59 [48 – 67] | 62 [53 – 68] |
| Male sex – no (%) | 128 (71.1) | 65 (78.3) | 63 (64.9) |
| Comorbidities – no (%) |  |  |  |
| Chronic respiratory disease (including asthma) | 20 (11.0) | 5 (6.0) | 15 (15.5) |
| Chronic heart failure (NYHA III or IV) | 6 (3.3) | 1 (1.2) | 5 (5.2) |
| Cardiovascular diseases (incl. hypertension) | 94 (51.9) | 38 (45.2) | 56 (57.7) |
| Diabetes requiring insulin | 15 (8.3) | 4 (4.8) | 11 (11.5) |
| Chronic kidney failure | 9 (5.0) | 1 (1.2) | 8 (8.2) |
| Liver cirrhosis (Child-Pugh B or more) | 1 (0.6) | 1 (1.2) | 0 (0.0) |
| Immunodepression | 21 (11.6) | 8 (9.5) | 13 (13.4) |
| BMI > 30 kg/m<sup>2</sup> – no (%) | 43 (27.4) | 20 (25.3) | 23 (29.5) |
| Treatment by ACEIs or ARBs – no (%) | 54 (30.3) | 26 (31.0) | 28 (29.8) |
| **COVID-19 data** |  |  |  |
| Time from symptom onset to admission, median (IQR) - days | 7 [5 – 10] | 8 [6 – 10] | 7 [4 – 10] |
| Confusion at admission – no (%) | 6 (3.4) | 0 (0.0) | 6 (6.2) |
| Respiratory rate, median (IQR) - /min | 26 [22 – 30] | 27 [24 – 32] | 26 [21 – 30] |
| Oxygen saturation (without oxygen), median (IQR) - % | 92 [89 – 94] | 92 [89 – 94] | 92 [90 – 94] |
| Oxygen flow at admission, median (IQR) – L/min | 2 [2 – 4] | 3 [2 – 4] | 2 [2 – 3] |
| Systolic blood pressure, median (IQR) – mmHg | 128<br>[113 – 143] | 124<br>[112 – 138] | 130<br>[116 – 146] |
| Lymphocyte count <500/mm<sup>3</sup> – no (%) | 15 (8.5) | 6 (7.2) | 9 (9.6) |
| C-reactive protein (CRP) > 40 mg/L – no (%) | 153 (86.0) | 76 (90.5) | 77 (81.9) |
| Proportion of lung affected on the CT scan > 50% – no (%) | 22 (16.9) | 14 (21.9) | 8 (12.1) |

### Propensity score model development

Propensity scores ranged from 0.11 to 0.90 and from 0.04 to 0.82 in the HCQ and no-HCQ groups, respectively, with 96.1% in the region of common support (**Supplementary data 1**). After applying IPTW, 15 of the 19 covariates in the planned propensity score had weighted standardised differences below 10% while 4 (confusion at admission, chronic kidney disease, heart failure [NYHA III or IV] and liver cirrhosis [Child-Pugh B or more]) exceeded the threshold (**Supplementary data 2**). These results were due to the absence of confusion at admission in the HCQ group (vs 6 patients with confusion in the no-HCQ group); only one patient had chronic kidney disease in the HCQ group (vs 8 in the control group); only one had heart failure (NYHA III or IV) (vs 5 patients in the no-HCQ group), and again only one had liver cirrhosis (Child-Pugh B or more) in the HCQ group (vs 0 in the no-HCQ group). These 4 variables were therefore not included in the final propensity score model (**Supplementary data 3**).

### Outcomes

In the IPTW analysis, 20.5% patients in the HCQ group were transferred to the ICU or died within 7 days, compared with 22.1% in the no-HCQ group (16 vs 21 events, RR 0.93, 95% CI 0.48–1.81) (**Table 2**). Sensitivity analyses provided consistent results with RR 0.88, 95% CI 0.49–1.57 in the unweighted sample; RR 0.90, 95% CI 0.45–1.77 in the trimmed analysis; and RR 0.95, 95% CI 0.47–1.93) when patients who received HCQ during their follow-up were excluded (**Supplementary data 4**). For the secondary outcomes, 2.8% of patients in the HCQ group died within 7 days, compared with 4.6% in the no-HCQ group (3 vs 4 events, RR 0.61, 95% CI 0.13–2.90). Similarly, 27.7% of the HCQ group and 24.1% of the no-HCQ group developed ARDS within 7 days (24 vs 23 events, RR 1.15, 95% CI 0.66–2.01]) (**Table 2**).

Results were similar in the subgroup of patients with better prognosis at admission (qSOFA <2) with RR 1.12, 95% CI 0.54−2.32 for the primary outcome, RR 1.09, 95% CI 0.15−7.76 for overall mortality, and RR 1.31, 95% CI 0.71−2.44 for ARDS.

**Table 2:** Primary and secondary outcomes. Weighted proportions, RRs and 95% CIs were obtained by inverse probability treatment weighting. *two missing data were removed from analysis. Abbreviations: CI, confidence interval; ICU, intensive care unit.

|  | HCQ (n=84) |  | No HCQ (n=97) |  | RR (95% IC) |
| --- | --- | --- | --- | --- | --- |
|  | Raw | Weighted proportion | Raw | Weighted proportion |  |
| **Death or transfer to ICU** | 16/84 (19.0) | 20.5 | 21/97 (21.6) | 22.1 | 0.93 (0.48 to 1.81) |
| **Day 7 mortality** | 3/84 (3.6) | 2.8 | 4/97 (4.1) | 4.6 | 0.61 (0.13 to 2.90) |
| **Occurrence of acute respiratory distress syndrome\*** | 24/84 (28.6) | 27.7 | 23/95 (24.2) | 24.1 | 1.15 (0.66 to 2.01) |

### Safety

Among the 84 patients receiving HCQ within the first 48 hours, 8 (9.5%) experienced ECG modifications requiring HCQ discontinuation at a median of 4 days (3-9) after it began, according to French national guidelines. Among them, 7 had a corrected QT interval (QTc) prolongation of more than 60 ms (including 1 with QTc > 500 ms). One patient who received no other medication that might interfere with cardiac conduction presented a first-degree atrioventricular block after 2 days of HCQ treatment. Of note, a patient in whom HCQ was initiated 5 days after admission (no-HCQ group) was transferred to the ICU 2 days afterwards, where he was prescribed lopinavir and ritonavir and developed left bundle branch block on day 8.

## Discussion

We report a comparative study that uses real-world data collected from routine care to assess the efficacy and safety of HCQ in a population of 181 patients hospitalised for COVID-19 hypoxemic pneumonia. We found that HCQ treatment at 600 mg/day added to standard of care was not associated with a reduction of admissions to ICUs or death 7 days after hospital admission, compared to standard of care alone. The rate of ARDS did not decrease either. Our population of patients hospitalised because they required oxygen is very similar to that reported by others, and the percentage of patients transferred to the ICU was similar to that reported in a Chinese cohort of 138 patients hospitalised for COVID-19 pneumonia.^16^ The clinical features of patients included were also consistent with other reports, with a predominance of men and of patients with cardiovascular comorbidities and/or obesity.^16,17^ Almost all patients had bilateral pneumonia, and 75% moderate or severe lung infiltrate.^18^ The patients in this study did not receive any other drug, in particular, potential confounders such as anti-viral and anti-inflammatory treatments, including steroids, before ICU admission.

The timing of antiviral treatment initiation may be critical in reducing SARS-Cov-2 viral load.^19^ Accordingly, in the recent lopinavir–ritonavir trial, a post-hoc subgroup analysis suggested that lopinavir–ritonavir could have a clinical benefit if started earlier than 12 days after the onset of symptoms.^20^ This was the case here, because (1) patients had a median time from symptom onset to inclusion of 7 days, (2) they were treated with HCQ as soon as possible (i.e. in the 48 hours after hospital admission), and (3) we found that viral RNA for SARS-CoV-2 was detectable for all patients at inclusion, showing active viral shedding. Previous reports^21^ indicate that HCQ should have been expected to show some antiviral efficacy. We did not check subsequent SARS-Cov-2 PCR in this study and therefore cannot reach a conclusion about its potential efficacy for decreasing viral shedding. Although this may appear to be a limitation, we used robust clinical outcomes here, i.e., death and ICU admission, which are substantially more clinically relevant.

COVID-19 pneumonia progression in the second week of illness is associated with a so-called “cytokine storm”,^17,22^ which is thought to be responsible for the clinical worsening of many patients. Most of the patients included in this study had an inflammatory syndrome defined by C-reactive-protein higher than 40 mg/l, which suggests that a cytokine storm syndrome had already begun.^23^ Drugs decreasing virus shedding may therefore be inadequate at this stage; this is why many anti-inflammatory drugs are currently being tested, such as tocilizumab, corticosteroids, and others. Despite the immunomodulatory properties of HCQ, which include regulation of the production of pro-inflammatory cytokines such as IL-2, IL-1, IL-6 and TNFα,^24^ and endosomal inhibition of toll-like receptors, which play a major role in innate immune response,25 this treatment showed no effectiveness in this specific population.

Finally, HCQ blocks the KCNH2-encoded hERG/Kv11.1 potassium channel and can potentially prolong the QTc, with potential severe consequences such as sudden cardiac death and cardiac arrhythmia.^26^ Besides QTc prolongations, we observed 2 other significant cardiologic events in this study, and the French national drug agency has reported 3 deaths potentially related to HCQ since its promotion to the public as a potential treatment for COVID-19. Although HCQ was considered safe in the context of SLE, these adverse events may be explained by the use of high-dose HCQ in elderly patients with renal impairment and frequent drug interactions. Accordingly, the negative clinical results of this study argue against the widespread use of HCQ in patients with COVID-19 pneumonia.

Our study has several limitations. First, although our aim was to emulate a target trial and we used robust statistical techniques for adjustment, treatment was not randomly assigned and potential unmeasured confounders may bias our results. Second, four potentially important prognostic variables could not be balanced in the PS model because none or only one patient in the HCQ group presented with these variables. Accordingly, caution is required in the interpretation of results, especially for overall mortality where only a limited number of events were observed. Third, we did not take a centre effect into account in the PS model because the number of patients treated with HCQ in centres was unbalanced (some centres treated all patients with HCQ, while others did not).

In conclusion, we found that HCQ did not significantly reduce admission to ICU or death at day 7 after hospital admission, or ARDS in hospitalised patients with hypoxemic pneumonia due to COVID-19. These results are of major importance and do not support the use of HCQ in patients hospitalised for a documented SARS-CoV-2 pneumonia.

## Data Availability

Data is available from authors upon reasonable request.

## Contributors

MM conceptualised the paper

MM, MR, AC, RP, CG, SG, RL, AS, FS, MM, MK, EC, BT, CM, PL, YS, MM, EP, NC, NR, VL, LM, XL, EA, NCC collected data.

VT, JD, EP, PR analysed data.

JP performed SARS-Cov2 RT-PCR

MM, VT, PR, BG, NCC interpreted the results and wrote the Article.

All authors contributed to revision of the final version of the manuscript.

## Acknowledgement

Ada Clarke, Moez Jallouli, Hicham Kardaoui, Kamil Chitour, Laetitia languille, Mouhamed Dieng, Jean-Daniel Lelièvre.

No financial support.

---

**Supplementary data 1:**
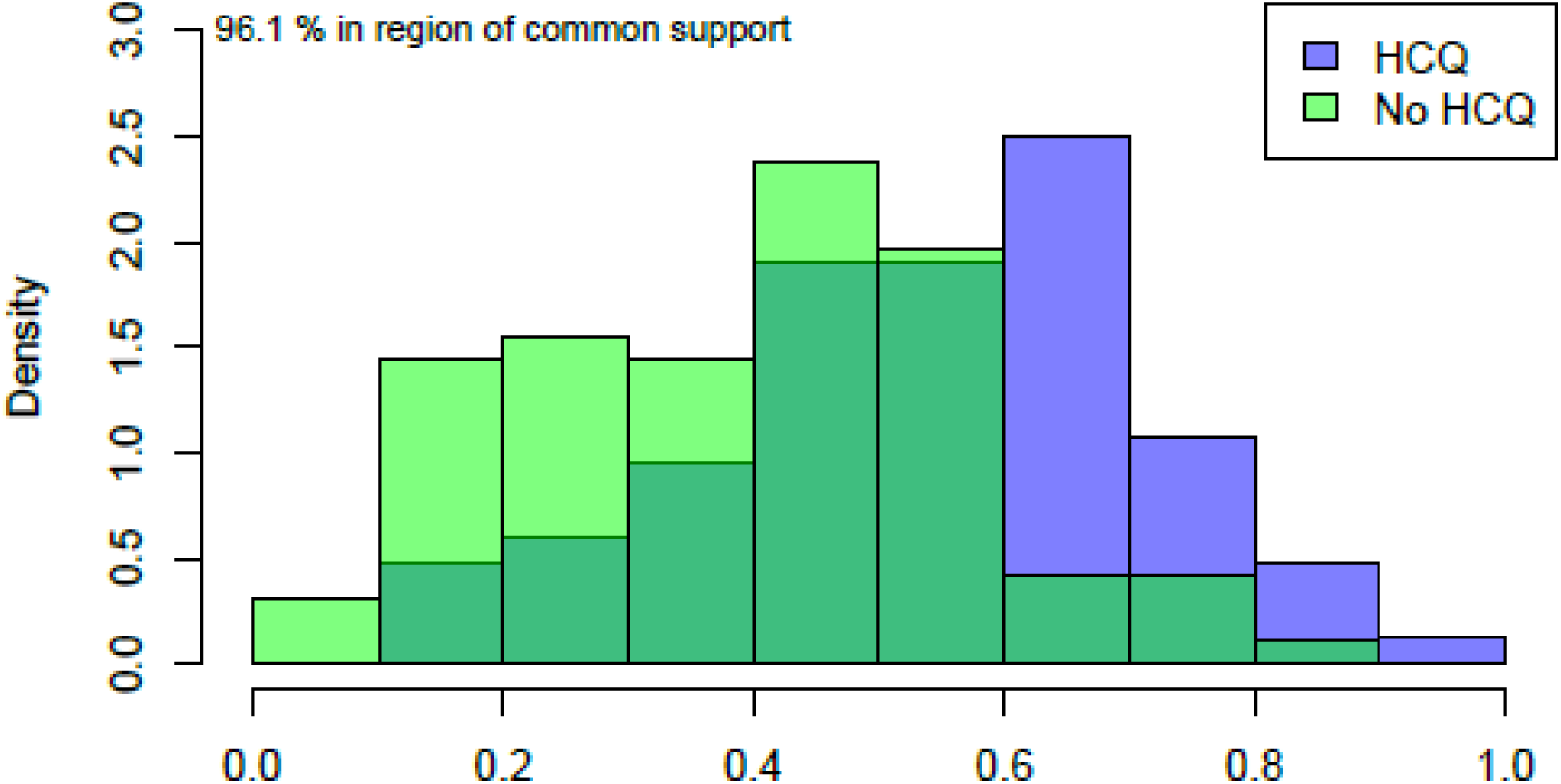
Propensity scores in the HCQ and no-HCQ groups.

**Supplementary data 2:**
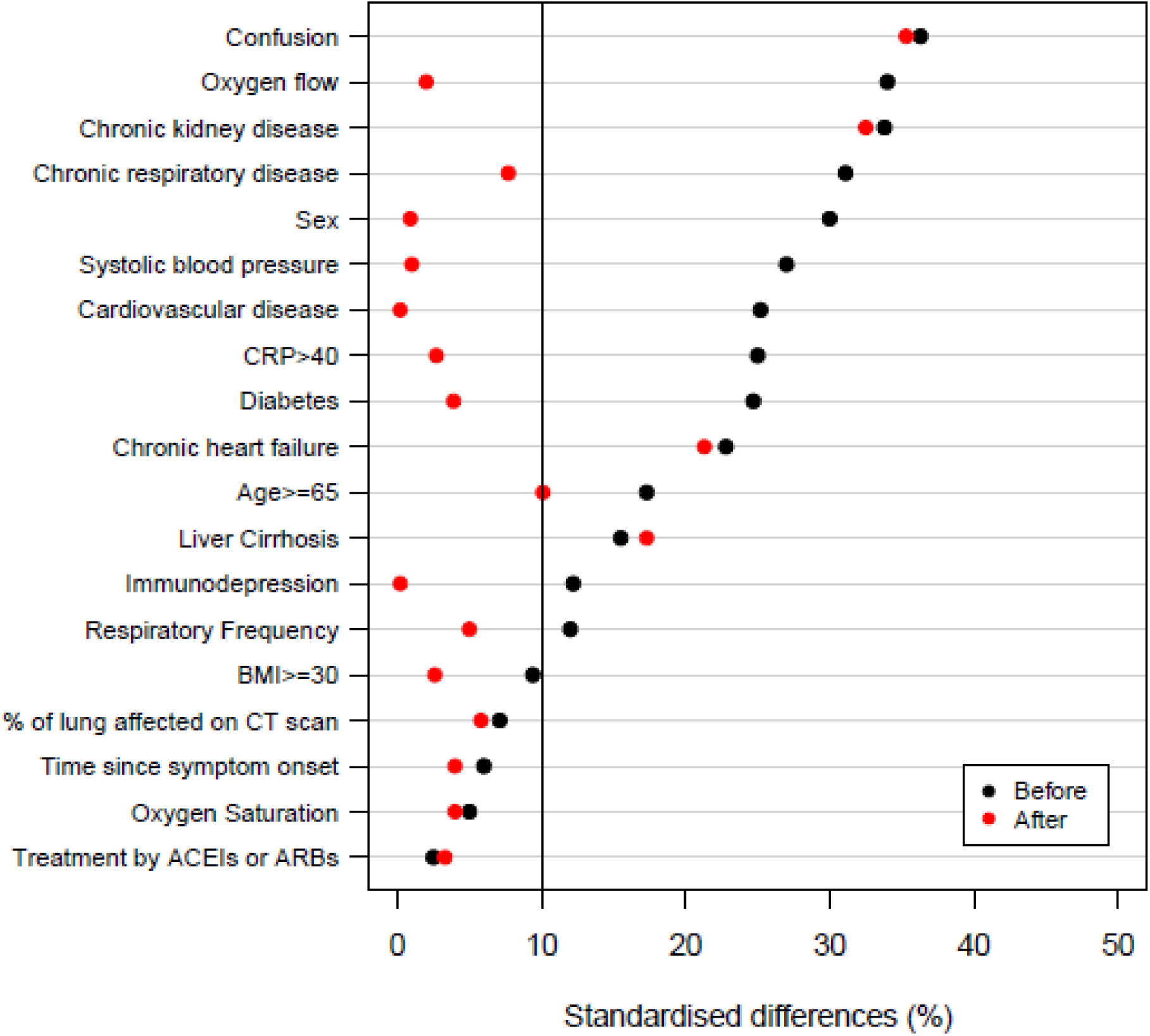
Standardised differences of variables used to generate the propensity score.

**Supplementary data 3:** Variables included in the final propensity score model.

| Variable | Type |
| --- | --- |
| Age | Binary ($\geq 65$ years old) |
| Gender | Binary (Male/Female) |
| Chronic respiratory insufficiency under oxygen therapy or asthma or cystic fibrosis or any chronic respiratory pathology likely to decompensate during a viral infection | Binary (Yes/No) |
| Cardiovascular disease: hypertension, stroke, coronary artery disease, cardiac surgery | Binary (Yes/No) |
| Insulin-dependent diabetes mellitus (DM), or DM with diabetic microangiopathy or macroangiopathy | Binary (Yes/No) |
| Treatment by immunosuppressive drugs (including anticancer chemotherapy) or an uncontrolled HIV infection or HIV infection with CD4 cell counts $< 200/\mu\text{l}$; or a haematological malignancy | Binary (Yes/No) |
| BMI | Binary ($\geq 30 \text{ kg/m}^2$ or not) |
| Treatment by angiotensin-converting enzyme inhibitors (ACEIs) or angiotensin receptor blockers (ARBs) | Binary (Yes/No) |
| Date of symptom onset | Continuous (days) |
| % of lung affected on the CT scan | Binary ($\geq 50\%$ or not) |
| Respiratory frequency | Continuous (Cycles/min) |
| Oxygen saturation without oxygen | Continuous (%) |
| Systolic blood pressure | Continuous (mmHg) |
| Need for oxygen and oxygen flow | Continuous (L/min) |
| C-Reactive Protein (CRP) ($\geq 40 \text{ mg/L}$ or not) | Binary ($\geq 40 \text{ mg/L}$ or not) |

**Supplementary data 4:** Sensitivity analyses*. Trimmed sample that was truncated at 10% of the extreme weights.

| Outcome | Analysis | HCQ | No-HCQ | RR (95% CI) |
| --- | --- | --- | --- | --- |
|  |  | No<br>(Weighted<br>proportion) | No<br>(Weighted<br>proportion) |  |
| Death or transfer to ICU by day 7 | Unweighted sample | 16/84 (19.0) | 21/97 (21.6) | 0.88 (0.49 – 1.57) |
|  | Trimmed sample* | 15/81 (20.3) | 21/93 (22.6) | 0.90 (0.45 – 1.77) |
|  | Exclusion of patients who received HCQ in the no-HCQ group | 16/84 (20.7) | 19/89 (21.8) | 0.95 (0.47 – 1.93) |
| Day 7 mortality | Unweighted sample | 3/84 (3.6) | 4/97 (4.1) | 0.87 (0.20 – 3.76) |
|  | Trimmed sample* | 3/81 (2.9) | 4/93 (4.8) | 0.61 (0.13 – 2.88) |
|  | Exclusion of patients who received HCQ in the no-HCQ group | 3/84 (3.6) | 4/89 (4.8) | 0.56 (0.12 – 2.67) |
| Acute respiratory distress syndrome by day 7 | Unweighted sample | 24/84 (28.6) | 23/95 (24.2) | 1.18 (0.72 – 1.93) |
|  | Trimmed sample* | 22/81 (27.0) | 23/91 (24.7) | 1.09 (0.62 – 1.94) |
|  | Exclusion of patients who received HCQ in the no-HCQ group | 24/84 (28.6) | 21/87 (23.9) | 1.19 (0.66 – 2.14) |

